# The Association between Marital Status and Suicide Risk Among Patients with Breast Cancer: A Population-Based sIPTW Competing Risk Analysis

**DOI:** 10.64898/2026.07.01.26357044

**Authors:** Xuehai Zou, Jingyi Shi

## Abstract

**Background:** Marital status at diagnosis may capture aspects of social support, but it is not a direct measure of support quality. This study examined its association with suicide mortality after breast cancer diagnosis while accounting for competing mortality and exploring variation by county-level income and rural-urban residence.

**Methods:** Adults of both sexes diagnosed with first primary breast cancer in SEER between 2000 and 2022 were studied using a baseline complete-case cohort. Marital status was classified as married/partnered or unmarried/non-partnered. Stabilized inverse probability of treatment weighting (sIPTW) was combined with multivariable Fine-Gray regression, treating non-suicide death as a competing event. Cox, landmark, exploratory subgroup, interaction, and unweighted treatment-adjusted sensitivity analyses were performed.

**Results:** The analytical cohort included 825,047 patients, of whom 40.7% were unmarried/non-partnered. All baseline standardized mean differences were below 0.01 after weighting. During 6,816,981.42 person-years, 506 suicide deaths occurred (7.42 per 100,000 person-years). Unmarried/non-partnered status was associated with higher suicide mortality in the primary Fine-Gray model (sHR 1.34, 95% CI 1.12-1.60) and Cox model (HR 1.45, 95% CI 1.21-1.73). Landmark sHRs at 1, 3, and 5 years were 1.39, 1.46, and 1.34, respectively. The treatment-adjusted sensitivity estimate was 1.26 (95% CI 0.99-1.60; P = 0.064). Income interaction analysis suggested heterogeneity (joint P = 0.042), but the Q4 conditional sHR was imprecise (1.06, 95% CI 0.80-1.39). There was no statistically significant rural-urban interaction (P = 0.488).

**Conclusions:** Unmarried/non-partnered status was associated with higher suicide mortality after breast cancer diagnosis, although the absolute rate was low. The observational design, sensitivity-analysis uncertainty, and exploratory income interaction warrant cautious interpretation and independent validation.

## Introduction

Breast cancer is the most diagnosed cancer among women worldwide and remains a major public health concern, with approximately 2.3 million new cases and 670,000 deaths reported globally in 2022 (1). As survival improves, the population of breast cancer survivors continues to expand, increasing attention toward long-term survivorship outcomes, including psychological well-being and suicide prevention (2, 3). However, breast cancer diagnosis and treatment remain associated with considerable psychological burden (2, 4). Breast cancer survivors commonly experience depression, anxiety, loneliness, altered body image, and reduced quality of life (4, 5, 6). A meta-analysis by Pilevarzadeh et al., including 72 studies from 30 countries, reported a pooled prevalence of depression of 32.2% among patients with breast cancer (4). In addition, long-term survivorship studies demonstrated that psychological distress and body-image disturbances may persist for years following surgery and treatment (5, 6). These psychological outcomes are clinically important because they may impair treatment adherence, reduce social functioning, and worsen survivorship experiences (4, 5, 6).

Suicide is an important concern among cancer survivors (7). A systematic review and meta-analysis reported suicidal ideation in approximately 10% of women with breast cancer and suicide attempts in approximately 2% (2). In a SEER study of female breast cancer survivors, Shi et al. observed 414 suicide deaths among 638,547 women and reported higher suicide risk than in the general population (8). This provides a rationale for examining social circumstances after diagnosis. The Social Buffering Framework proposes that support can moderate responses to major stressors (9). Marital status is frequently used as a sociodemographic proxy for interpersonal support, although it does not directly measure the support received (9, 10, 11, 12, 13). Prior breast cancer studies have primarily examined the relationship between marital status and overall or cancer-specific survival (10, 11, 12, 13). These outcomes differ from suicide mortality and do not by themselves establish the association of interest here.

Socioeconomic and geographic circumstances may also shape breast cancer presentation, care, and survivorship (14, 15, 16, 17). Ayub et al. studied 815,220 breast cancer cases in SEER from 2004 to 2020 and reported differences in clinical presentation and treatment patterns across rural-urban and income groups (14). That study did not report a suicide mortality rate or a marital-status effect on suicide. Its findings therefore provide context for potential geographic and socioeconomic differences in care, rather than direct evidence about suicide mortality. Research on deprivation, Indigenous populations, and access to screening further illustrates the relevance of social and geographic context, while addressing different populations and outcomes (15, 16, 17).

Although competing-risk analyses of suicide among breast cancer survivors have been published (8), evidence on whether the marital-status association varies by area-level income or rural-urban residence remains limited. We therefore examined marital status and suicide mortality using an sIPTW-weighted multivariable Fine-Gray model, with non-suicide deaths treated as competing events. Income and rural-urban interactions were evaluated as exploratory extensions of the primary association analysis.

## Methods

### Data Source and Study Population

Data were obtained from the SEER Research Data database, 17 Registries, November 2024 submission, covering diagnoses in 2000-2022 and linked county attributes (18). Adult patients of both sexes with microscopically confirmed first primary breast cancer were identified using SEER*Stat. The clinical selection excluded diagnoses based only on autopsy or death certificate. The exported dataset contained 1,105,193 records. Excluding 12,131 records with survival shorter than 1 month left 1,093,062 records. The minimum follow-up requirement defined the analysis population and avoided zero recorded follow-up; it was not a method for removing competing mortality (Figure 1).

**Figure 1:**
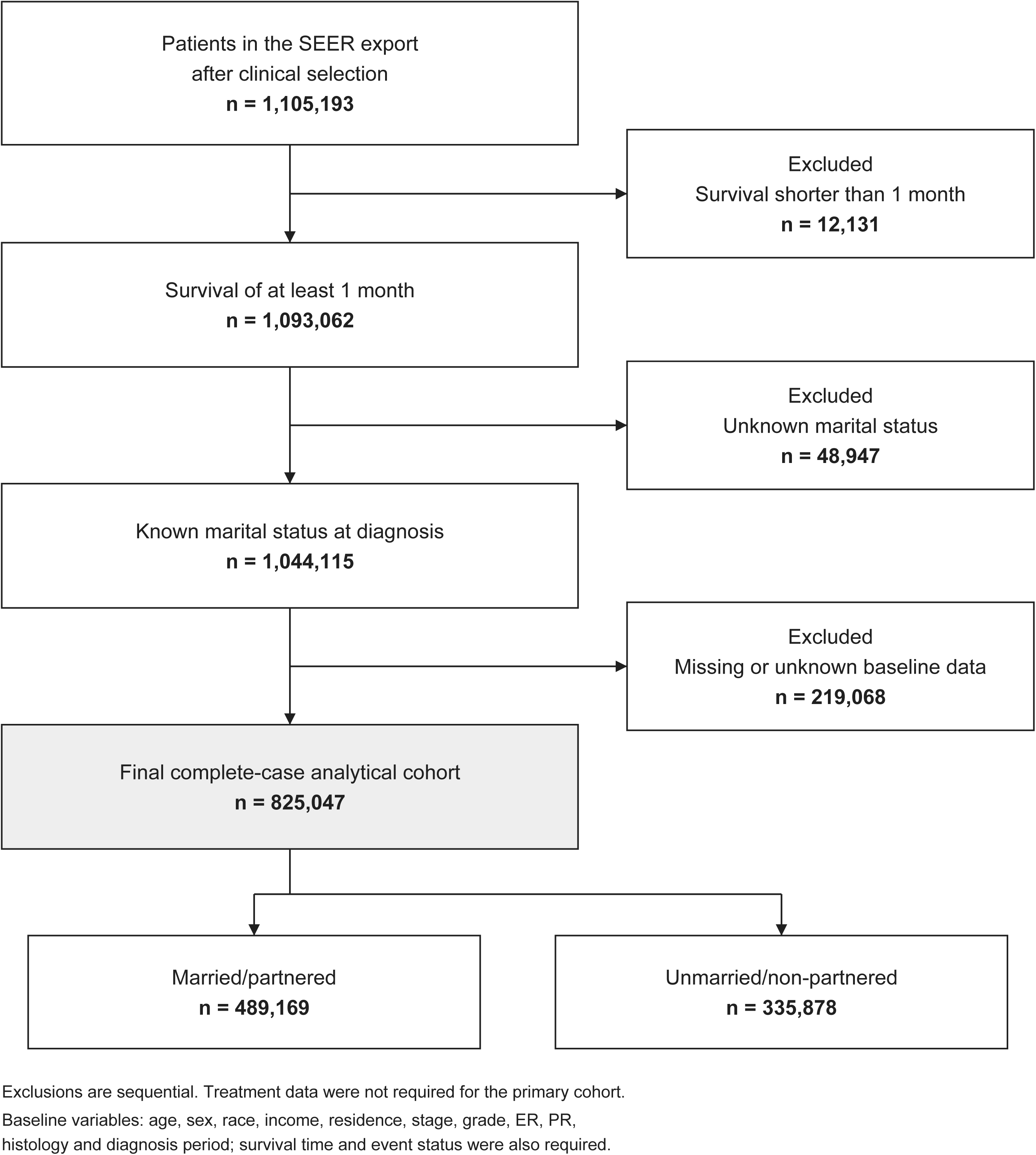
Selection process of adult patients of both sexes diagnosed with primary breast cancer. The SEER export contained 1,105,193 records after clinical selection. Excluding 12,131 with survival <1 month left 1,093,062; excluding 48,947 with unknown marital status left 1,044,115; excluding 219,068 with missing/unknown baseline data yielded 825,047 complete cases. Each number refers to a successive step rather than a competing definition of the final cohort.

Married/partnered status included “Married (including common law)” and “Unmarried or Domestic Partner.” Unmarried/non-partnered status included “Single (never married),” “Widowed,” “Divorced,” and “Separated.” Exclusion of 48,947 records with unknown marital status left 1,044,115 records. A further 219,068 records were excluded for missing or unknown baseline data, yielding 825,047 complete cases (19).

Required fields included age, sex, race, income category, rural-urban residence, stage, grade, ER status, PR status, histology, diagnosis period, survival time, and event status. Treatment information was not required for entry to the primary cohort. The combined No/Unknown categories for chemotherapy and radiation were retained in treatment-related analyses. Marital status was measured at diagnosis and was treated as a limited proxy for interpersonal circumstances; heterogeneous unmarried categories were combined, and support quality was not measured.

### Covariates

Baseline covariates were age (<65 or ≥65 years), sex (female or male), race, county-level median household income, rural-urban residence, tumor stage, tumor grade, estrogen receptor (ER) status, progesterone receptor (PR) status, and histology. Income was inflation-adjusted to 2023 and grouped using fixed cutoffs: Q1, <$55,000; Q2, $55,000-$69,999; Q3, $70,000-$84,999; and Q4, ≥$85,000. These were prespecified income categories, not empirical quartiles of the analytical cohort. Detailed descriptive categories included White, Black, Asian or Pacific Islander, and American Indian/Alaska Native race; localized, regional, and distant stage; and Grades I-IV. ER and PR were positive or negative, and included histologies were infiltrating ductal, infiltrating lobular, and mixed ductal/lobular carcinoma. Surgery was recoded as breast-conserving surgery, mastectomy, no surgery, or unknown. Chemotherapy and radiation were coded as Yes or the SEER combined No/Unknown category.

### Outcome and Estimand Assessment

The primary outcome was suicide mortality, defined using SEER cause-of-death recode variables. Follow-up time was calculated from diagnosis to suicide death, death from other causes, or end of follow-up.

The primary estimand was the association between marital status at diagnosis and subsequent suicide mortality after accounting for measured demographic, socioeconomic, and tumor-related characteristics at diagnosis. Because some clinical characteristics, such as stage at diagnosis, may partly reflect pathways related to healthcare access or social support, the estimates should not be interpreted as the total causal effect of marital status.

### Stabilized Inverse Probability of Treatment Weighting (sIPTW)

A logistic regression model estimated e(X), the probability of unmarried/non-partnered status conditional on the same baseline covariates used in the primary outcome model: age, sex, binary race, income category, rural-urban residence, binary stage, binary grade, ER, PR, and histology. If p denotes the observed proportion unmarried/non-partnered, stabilized weights were p/e(X) for unmarried/non-partnered patients and (1 − p)/(1 − e(X)) for married/partnered patients (20). The code checked model convergence, finite propensity scores strictly between 0 and 1, and finite positive weights. No weight truncation or trimming was applied. Weight distributions and effective sample size, calculated as (sum of weights) squared divided by the sum of squared weights, were examined. Baseline balance was assessed using standardized mean differences (SMDs), with an absolute SMD below 0.10 considered adequate (21). These empirical checks do not establish the absence of unmeasured confounding.

### Fine-Gray Competing-Risk Analysis and Cox Hazard Ratio Regression

The primary model was an sIPTW-weighted multivariable Fine-Gray regression for suicide mortality (22). Event status was coded as 0 for alive/censored, 1 for suicide death, and 2 for non-suicide death. The survival package finegray function expanded the data into start-stop intervals. Final analysis weights were the product of the Fine-Gray expansion weight (fgwt) and each patient’s baseline sIPTW. The model used Surv(fgstart, fgstop, fgstatus), included marital status and the baseline covariates listed above, and estimated robust standard errors clustered on a stable patient identifier. Estimates were reported as subdistribution hazard ratios (sHRs) with 95% confidence intervals; they describe relative subdistribution hazards rather than absolute risk differences (23).

For the propensity score and primary, Cox, and landmark models, race was coded as White versus non-White, stage as localized versus regional/distant, and grade as lower (Grades I-II) versus higher (Grades III-IV). Married/partnered, age <65 years, female, White, Q1 income, urban residence, localized stage, lower grade, ER positive, PR positive, and infiltrating ductal histology were the reference categories. Treatment variables were omitted from the primary models because they may lie on post-diagnosis pathways linking social circumstances and outcomes.

An sIPTW-weighted cause-specific Cox model with the same baseline covariates and robust variance estimation was also fitted, censoring non-suicide deaths at their recorded death times. Its proportional hazards assumption was evaluated using Schoenfeld residual tests (24, 25). No formal time-interaction test of the Fine-Gray proportional subdistribution hazards assumption was performed. Model-based cumulative incidence curves were predicted for two profiles differing only in marital status, with all other covariates set to their reference categories; these are reference-profile predictions, not population-standardized cumulative incidence estimates (Supplementary Figure 2).

### Landmark Analyses

Landmark analyses were performed at 12, 36, and 60 months after diagnosis among patients whose recorded survival exceeded the corresponding landmark, so that they were still alive and under observation immediately after that time (26). Follow-up was reset by subtracting the landmark month. Subsequent suicide was the event of interest and subsequent non-suicide death was a competing event. Fine-Gray expansion was repeated within each landmark cohort. The original baseline sIPTW was retained without re-estimation and multiplied by the landmark-specific expansion weight. Models included the same baseline covariates and used patient-clustered robust variance. These estimates are conditional associations among successive survivor cohorts, not estimates of an effect in the original population.

### Subgroup Analysis

Exploratory subgroup analyses used the baseline complete-case cohort and strata defined by age, detailed race, diagnosis period (2000-2010 or 2011-2022), stage, grade, surgery, chemotherapy, and radiation. Within each stratum, Fine-Gray expansion was repeated and the expansion weights were multiplied by the original baseline sIPTW. Models adjusted for the remaining baseline covariates, omitting the stratification variable, its corresponding binary recode, and any invariant covariates. Strata with fewer than 200 patients, fewer than 10 suicide events, or only one marital-status group were not estimated. Fits with warnings, errors, or nonfinite estimates were recorded as not estimable. Unknown surgery was excluded from surgery-specific strata; chemotherapy and radiation No/Unknown categories were retained.

Subgroup analyses were conducted as exploratory analyses to assess the consistency of the marital-status association across selected demographic, clinical, and treatment-related strata. Because of the limited number of suicide deaths, subgroup estimates were interpreted cautiously and were not considered evidence of effect modification unless supported by a formal interaction test.

### Interaction Analysis

Two separate interaction models extended the primary model: marital status × income category plus the other baseline covariates, and marital status × rural-urban residence plus the other baseline covariates. Each contained both main effects and their product terms, used start-stop Fine-Gray expanded data, applied fgwt × sIPTW, and used patient-clustered robust variance. Married/partnered, Q1, and urban residence were the exposure and moderator reference levels. Joint Wald tests evaluated the three income interaction coefficients and the single rural interaction coefficient (27). Within each moderator level, the conditional marital-status log-sHR was the sum of the marital main-effect coefficient and the applicable interaction coefficient. Its standard error was calculated from the full robust covariance matrix, including the covariance between those coefficients, before exponentiation to obtain the sHR and confidence interval.

### Sensitivity Analyses

An unweighted treatment-adjusted Fine-Gray sensitivity model started from the same baseline complete-case cohort. Records with missing treatment data or a standalone Unknown treatment category were excluded; the observed additional exclusions arose from surgery classified as unknown. The combined chemotherapy and radiation No/Unknown categories were retained. The model included all primary baseline covariates, including sex, plus surgery, chemotherapy, and radiation. Fine-Gray expansion weights were used without sIPTW, and robust standard errors were clustered by patient. Because both the analyzed sample and the weighting/adjustment specification differed from the primary model, a change in the estimate cannot be attributed to treatment adjustment alone.

### Statistical Analysis

Analyses were implemented in R 4.5.2 using packages including survival, tidyverse, survey, cobalt, gtsummary, broom, gt, and patchwork. The workflow set the random seed to 20260513, checked required fields and packages, and included sessionInfo output for reproducibility. Missing and unknown baseline covariates were handled by complete-case exclusion, without multiple imputation. Regression inference used robust standard errors; interactions were evaluated by joint Wald tests. Tests were two-sided, with P < 0.05 used as a descriptive significance threshold. Subgroup and interaction analyses were exploratory, with no multiplicity adjustment, and were interpreted separately from the primary analysis.

## Results

### Baseline Characteristics

The export contained 1,105,193 records, of which 1,093,062 had survival of at least 1 month and 1,044,115 had known marital status. After baseline complete-case exclusions, 825,047 patients remained: 489,169 married/partnered and 335,878 unmarried/non-partnered (40.7%; Figure 1). Both baseline tables describe this final cohort. Mean age was 58.2 years (SD 12.3) in the married/partnered group and 63.1 years (SD 14.5) in the unmarried/non-partnered group. The corresponding proportions aged <65 years were 68% and 52%, and the proportions classified as White were 82% and 77% (Table 1; Supplementary Table 1).

**Table 1:**
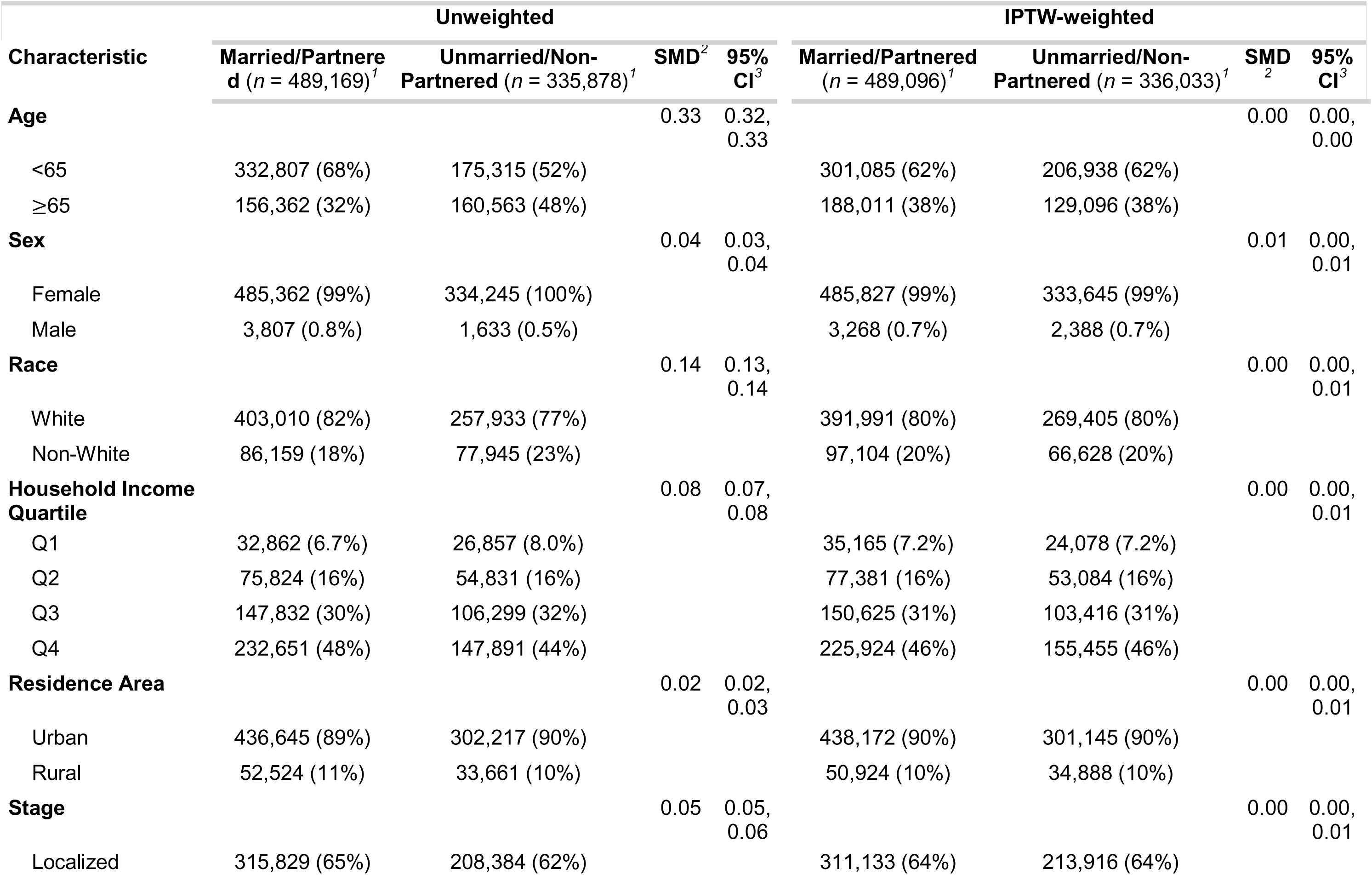
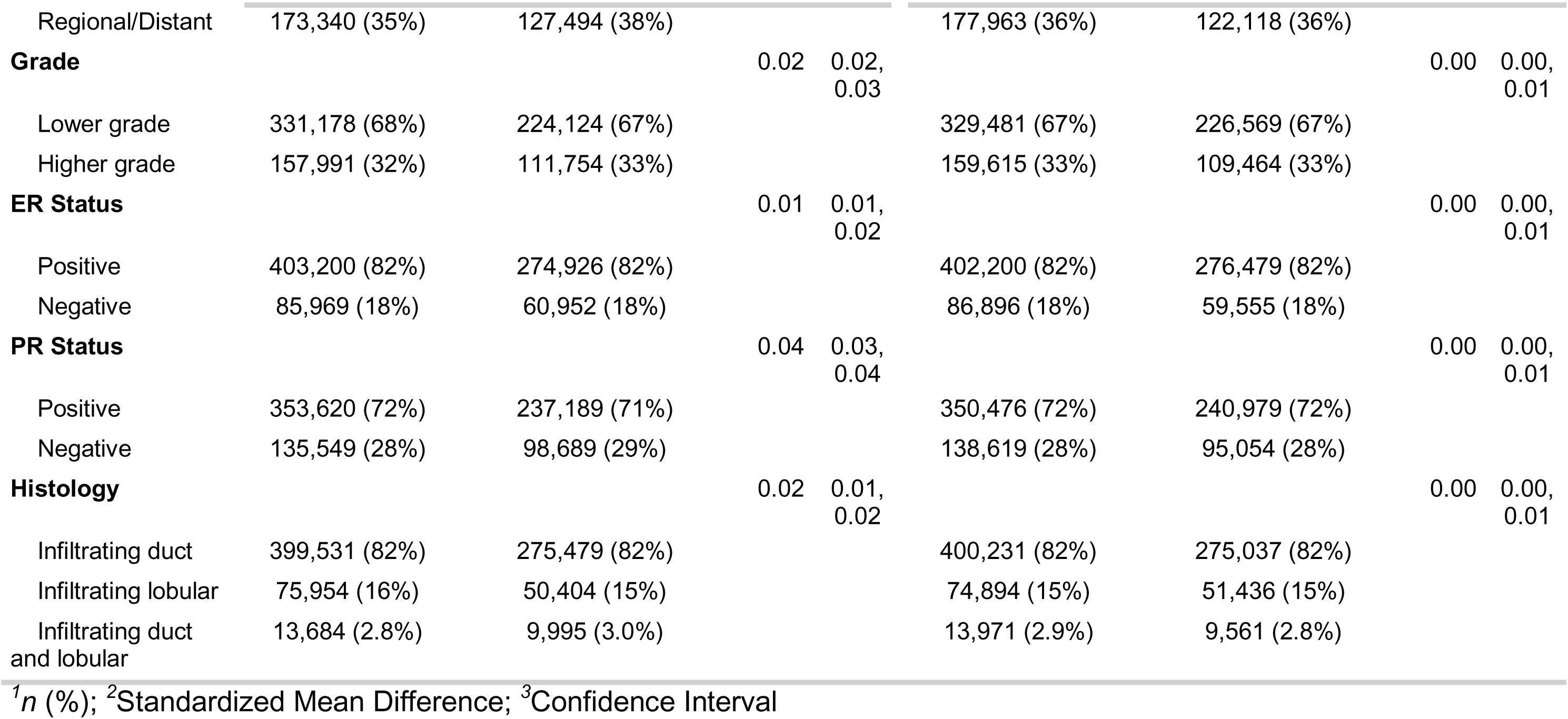
Patient Characteristics Pre- and Post-Weighting by Marital Status. *^1^n (%); ^2^Standardized Mean Difference; ^3^Confidence Interval* Abbreviations: CI, confidence interval; ER, estrogen receptor; IPTW, inverse probability of treatment weighting; PR, progesterone receptor; SMD, standardized mean difference. Both unweighted and weighted columns refer to the 825,047-patient baseline complete-case cohort. Values are n (%) or weighted n (%), respectively; weighted counts are not additional patients. Marital groups are married/partnered and unmarried/non-partnered. Baseline variables and reference categories are defined in the Methods. SMDs and their reported confidence intervals describe balance before and after stabilized weighting; the prespecified absolute SMD threshold was 0.10. SEER cause-of-death recode identified suicide, and non-suicide deaths were competing events. Income categories Q1-Q4 use fixed monetary cutoffs rather than sample quartiles.

### Stabilized Inverse Probability of Treatment Weighting (sIPTW)

Before weighting, the largest SMDs were observed for age (0.328) and race (0.139). After sIPTW, all SMDs were below 0.01; the largest was 0.00536 for sex (Table 1; Supplementary Figure 1). Stabilized weights ranged from 0.571 to 2.698, with a median of 0.926 and 99th percentile of 1.506. The effective sample size was 793,642.7, compared with 825,047 observed patients.

### Association Between Marital Status and Suicide Mortality

During 6,816,981.42 person-years of observation, 506 suicide deaths occurred. Median observed follow-up was 85 months (interquartile range, 38-151 months). The crude suicide mortality rate was 7.42 per 100,000 person-years overall, 6.71 among married/partnered patients (287 deaths), and 8.62 among unmarried/non-partnered patients (219 deaths). In the primary model, unmarried/non-partnered status was associated with a higher subdistribution hazard of suicide (sHR 1.34, 95% CI 1.12-1.60; P = 0.0014; Figure 2 and Table 2). Reference-profile cumulative incidence curves are presented in Supplementary Figure 2.

**Figure 2:**
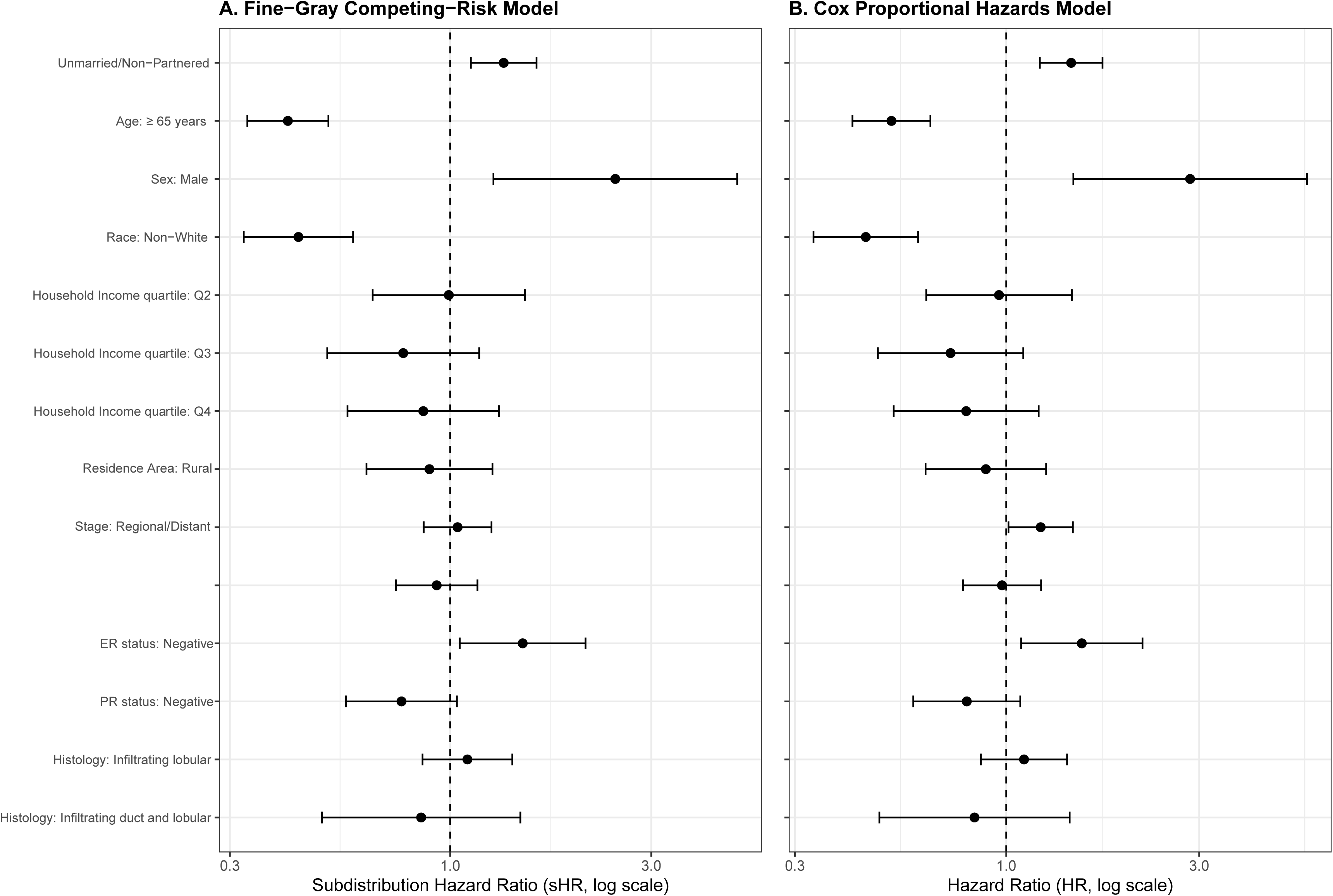
Combined Fine−Gray competing−risk model and Cox proportional hazards model results. Panel A presents sIPTW-weighted multivariable Fine-Gray sHRs and panel B presents sIPTW-weighted cause-specific Cox HRs, with 95% CIs and the same baseline covariates. Non-suicide death is a competing event in panel A and is censored at death in panel B. The reference line is 1.0. Cox diagnostics indicated non-proportional hazards for stage and grade. Reference categories are given in the Methods.

**Table 2:**
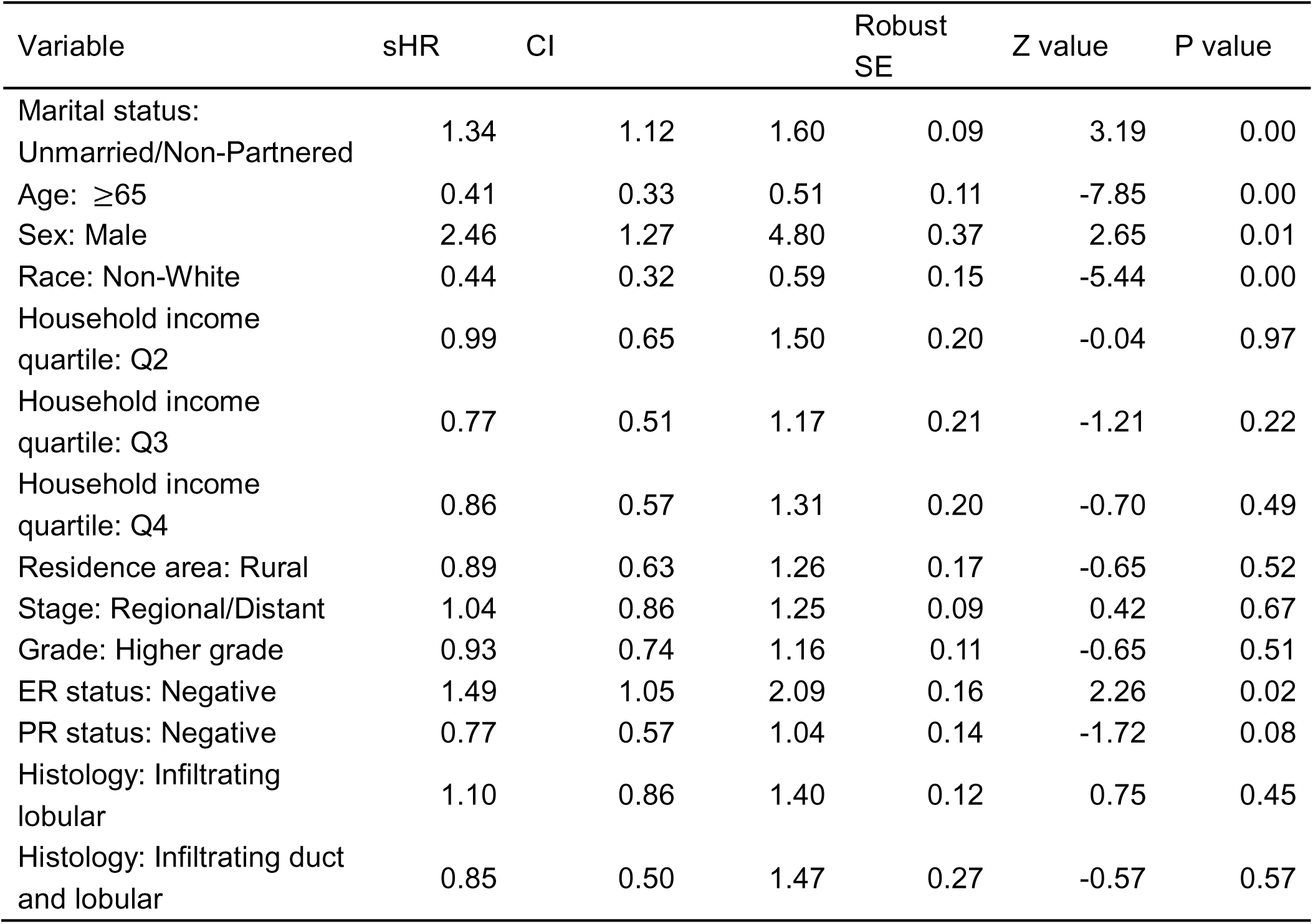
Fine-Gray competing risk model results. Abbreviations: CI, confidence interval; sHR, subdistribution hazard ratio; SE, standard error. The primary sIPTW-weighted multivariable Fine-Gray model used the baseline complete-case cohort and included marital status, age, sex, race, income category, rural-urban residence, stage, grade, ER, PR, and histology. Final weights were fgwt × sIPTW. Reported sHRs, 95% CIs, and P values use robust variance clustered by patient. Reference categories are married/partnered, <65 years, female, White, Q1, urban, localized, lower grade, ER positive, PR positive, and infiltrating ductal histology. Treatment was not included in this model.

In the cause-specific Cox analysis, the marital-status HR was 1.45 (95% CI 1.21-1.73; P < 0.001). The global Schoenfeld test gave P = 0.426, while stage (P = 0.0197) and grade (P = 0.0463) showed evidence of non-proportional hazards. Thus, the Cox result should not be interpreted as confirming proportional hazards for every covariate; no time-varying coefficient correction was applied.

At the 1-year landmark, 754,044 patients remained under observation and 438 subsequent suicide deaths occurred; the marital-status sHR was 1.39 (95% CI 1.15-1.69; P < 0.001). At 3 years, 626,851 patients and 324 subsequent suicide deaths contributed to an sHR of 1.46 (95% CI 1.17-1.83; P < 0.001). At 5 years, 517,597 patients and 231 subsequent suicide deaths contributed to an sHR of 1.34 (95% CI 1.03-1.75; P = 0.0313). These are conditional associations in the respective survivor cohorts (Figure 3; Table 3).

**Figure 3:**
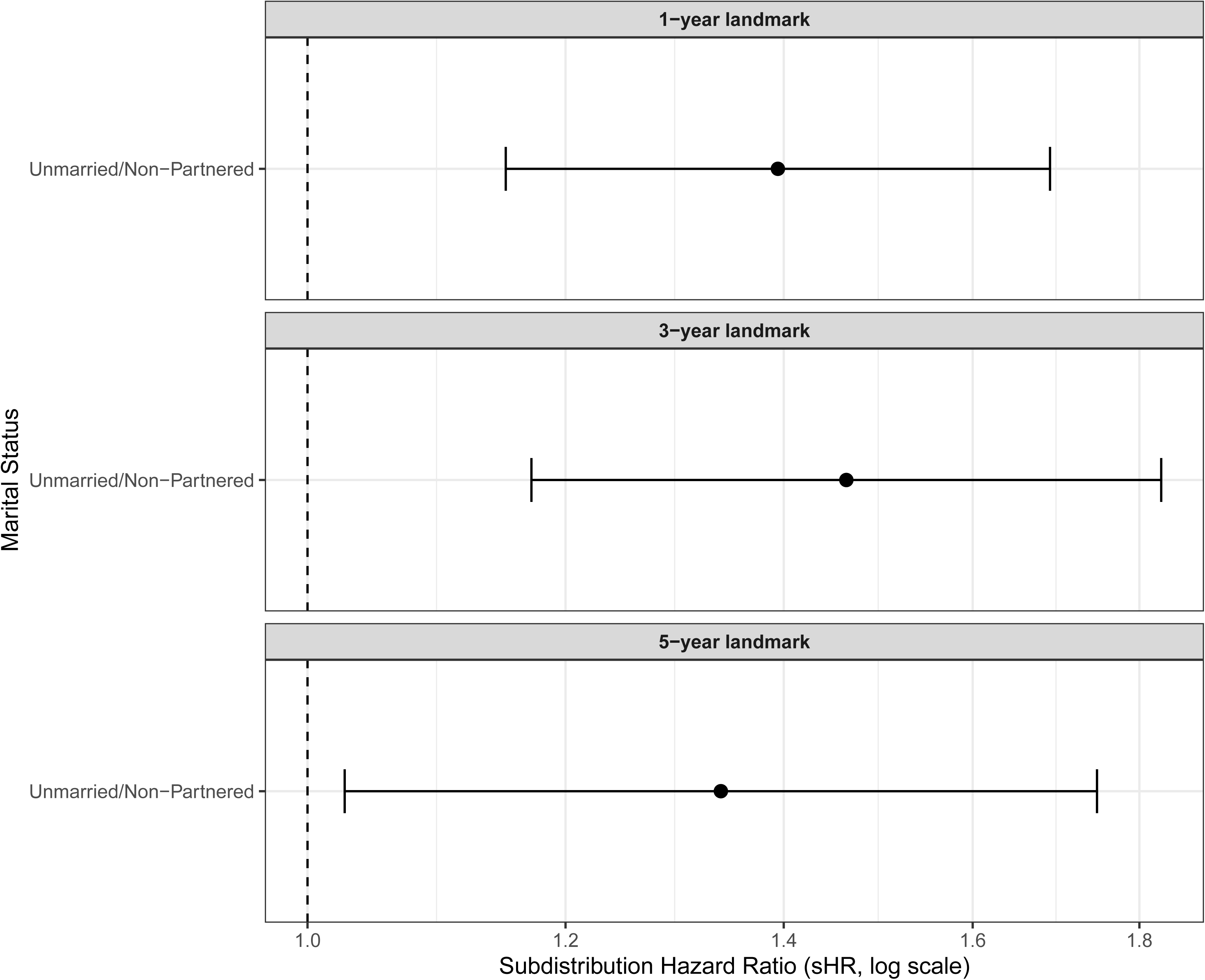
Landmark Fine-Gray analysis of marital status and suicide mortality. Conditional marital-status associations at 12, 36, and 60 months after diagnosis are shown as sHRs with 95% CIs. Patients had recorded survival beyond each landmark; follow-up was reset and baseline sIPTW retained. Married/partnered is the reference. Cohort sizes were 754,044, 626,851, and 517,597, with 438, 324, and 231 subsequent suicide deaths, respectively. The estimates apply to successive survivor cohorts.

**Table 3.** Association Between Marital Status and Suicide Mortality Among Breast Cancer Survivors Stratified by Post-Diagnosis Survival Intervals (Landmark Analysis). Abbreviations: CI, confidence interval; sHR, subdistribution hazard ratio; SE, standard error. Landmark cohorts comprised patients with recorded survival beyond 12, 36, or 60 months. Follow-up was reset at each landmark; the original baseline sIPTW was retained and multiplied by newly generated Fine-Gray weights. Models adjusted for the primary baseline covariates and used patient-clustered robust variance. Married/partnered is the marital reference. Sample sizes and subsequent suicide events were 754,044/438, 626,851/324, and 517,597/231, respectively. Dashes identify reference or non-estimated parameters, not zero effects.

| Landmark Interval | Marital Status | Patients at Risk (n)* | Subsequent Suicide Events (n) | Primary Competing-Risk Model: sHR (95% CI) | P-Value | Sensitivity Analysis: Cox HR (95% CI) | P-Value |
| --- | --- | --- | --- | --- | --- | --- | --- |
| 1-Year Landmark | Married/Partnered | 451,703 | 246 | — | — | — | — |
| (Surviving ≥12 months) | Unmarried/Non-Partnered | 302,341 | 192 | 1.39 (1.15–1.69) | <0.001 | 1.50 (1.24–1.81) | <0.001 |
| 3-Year Landmark | Married/Partnered | 382,667 | 182 | — | — | — | — |
| (Surviving ≥36 months) | Unmarried/Non-Partnered | 244,184 | 142 | 1.46 (1.17–1.83) | <0.001 | 1.55 (1.25–1.94) | 0.031 |
| 5-Year Landmark | Married/Partnered | 321,238 | 138 | — | — | — | — |
| (Surviving ≥60 months) | Unmarried/Non-Partnered | 196,359 | 93 | 1.34 (1.03–1.75) | 0.031 | 1.41 (1.08–1.84) | 0.011 |

### Association Between Socioeconomic Status and Suicide Mortality

In the primary Fine-Gray model, income categories Q2-Q4 were not statistically significantly associated with suicide mortality compared with Q1 (sHRs 0.77-0.99; all P > 0.20). Rural versus urban residence also showed no statistically significant association (sHR 0.89, 95% CI 0.63-1.26; P = 0.517). These main-model estimates are distinct from the conditional marital-status associations in the interaction models.

### Association Between Other Covariates and Suicide Mortality

Age ≥65 years, compared with <65 years, was associated with a lower subdistribution hazard of suicide (sHR 0.41, 95% CI 0.33-0.51). Non-White versus White race was also associated with a lower subdistribution hazard (sHR 0.44, 95% CI 0.32-0.59). Male versus female sex (sHR 2.46, 95% CI 1.27-4.80) and ER-negative versus ER-positive tumors (sHR 1.49, 95% CI 1.05-2.09) were associated with higher subdistribution hazards (Table 2).

There were no statistically significant associations for regional/distant versus localized stage (sHR 1.04, 95% CI 0.86-1.25), higher versus lower grade (sHR 0.93, 95% CI 0.74-1.16), or PR-negative versus PR-positive status (sHR 0.77, 95% CI 0.57-1.04). Compared with infiltrating ductal histology, the estimates were 1.10 (95% CI 0.86-1.40) for infiltrating lobular and 0.85 (95% CI 0.50-1.47) for mixed ductal/lobular histology.

### Interaction analyses

Income interaction analysis suggested heterogeneity in the marital-status association (joint Wald chi-square = 8.18, 3 df; P = 0.0424). Conditional sHRs for unmarried/non-partnered versus married/partnered status were 2.74 (95% CI 1.43-5.26; P = 0.0024) in Q1, 1.52 (0.99-2.31; P = 0.0535) in Q2, 1.48 (1.07-2.05; P = 0.0178) in Q3, and 1.06 (0.80-1.39; P = 0.6859) in Q4. The Q4 multiplicative interaction ratio relative to Q1 was 0.386 (95% CI 0.191-0.782; P = 0.0082). This suggests possible attenuation in Q4 but does not establish absence of an association within that category. The rural-urban interaction was not statistically significant (joint P = 0.4877); conditional marital-status sHRs were 1.31 (95% CI 1.08-1.58) in urban and 1.60 (0.94-2.75) in rural areas (Figure 4; Table 4).

**Figure 4:**
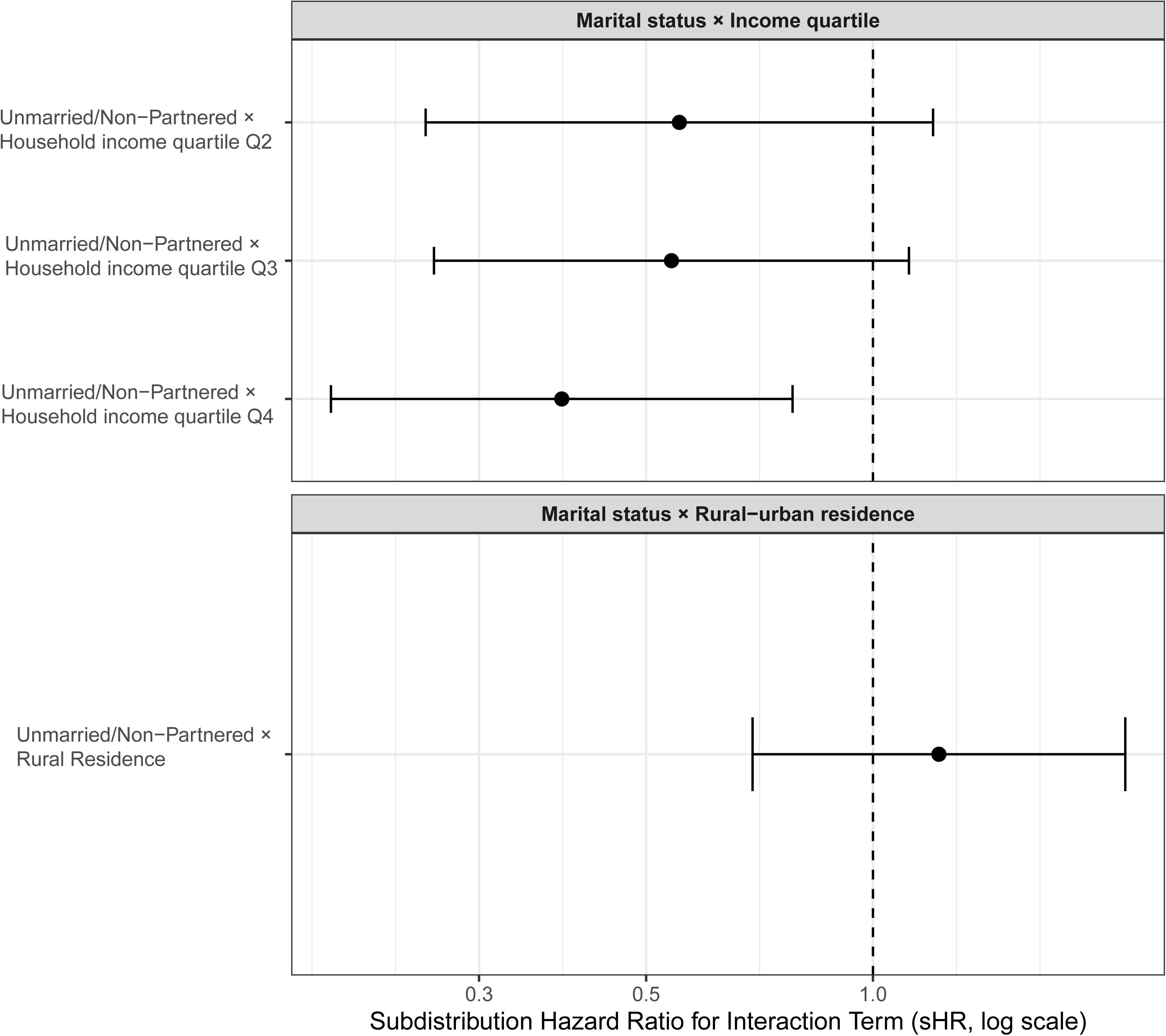
Interaction analysis of covariates and marital status on suicide risk. The plotted product-term estimates are multiplicative interaction ratios, not the stratum-specific conditional marital-status sHRs reported in Table 4. Married/partnered is the exposure reference; Q1 and urban residence are moderator reference levels. All 95% CIs use robust variance. Joint interaction P values are 0.0424 for income (3 df) and 0.4877 for rural-urban residence (1 df). The reference line is 1.0.

**Table 4:**
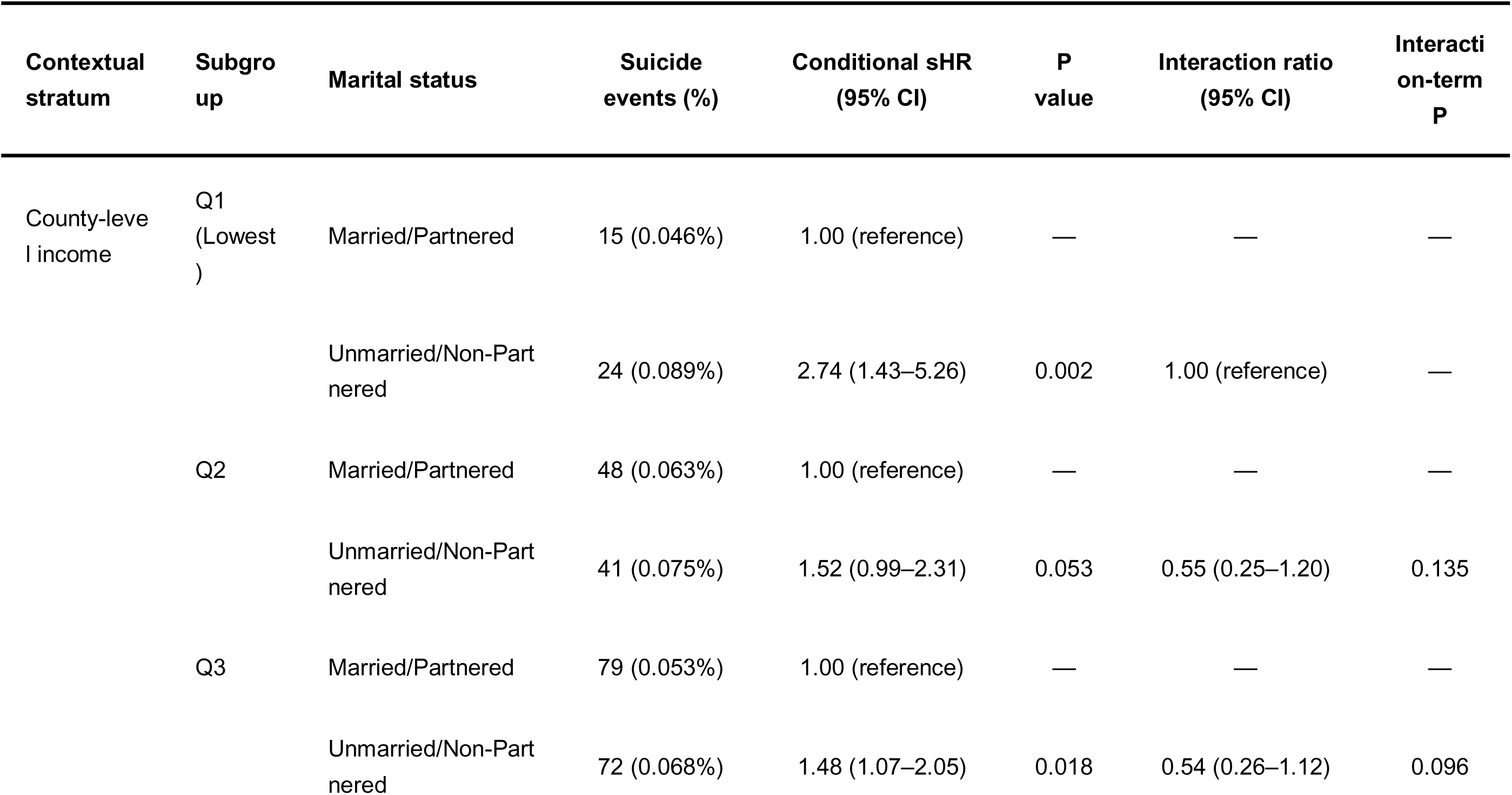
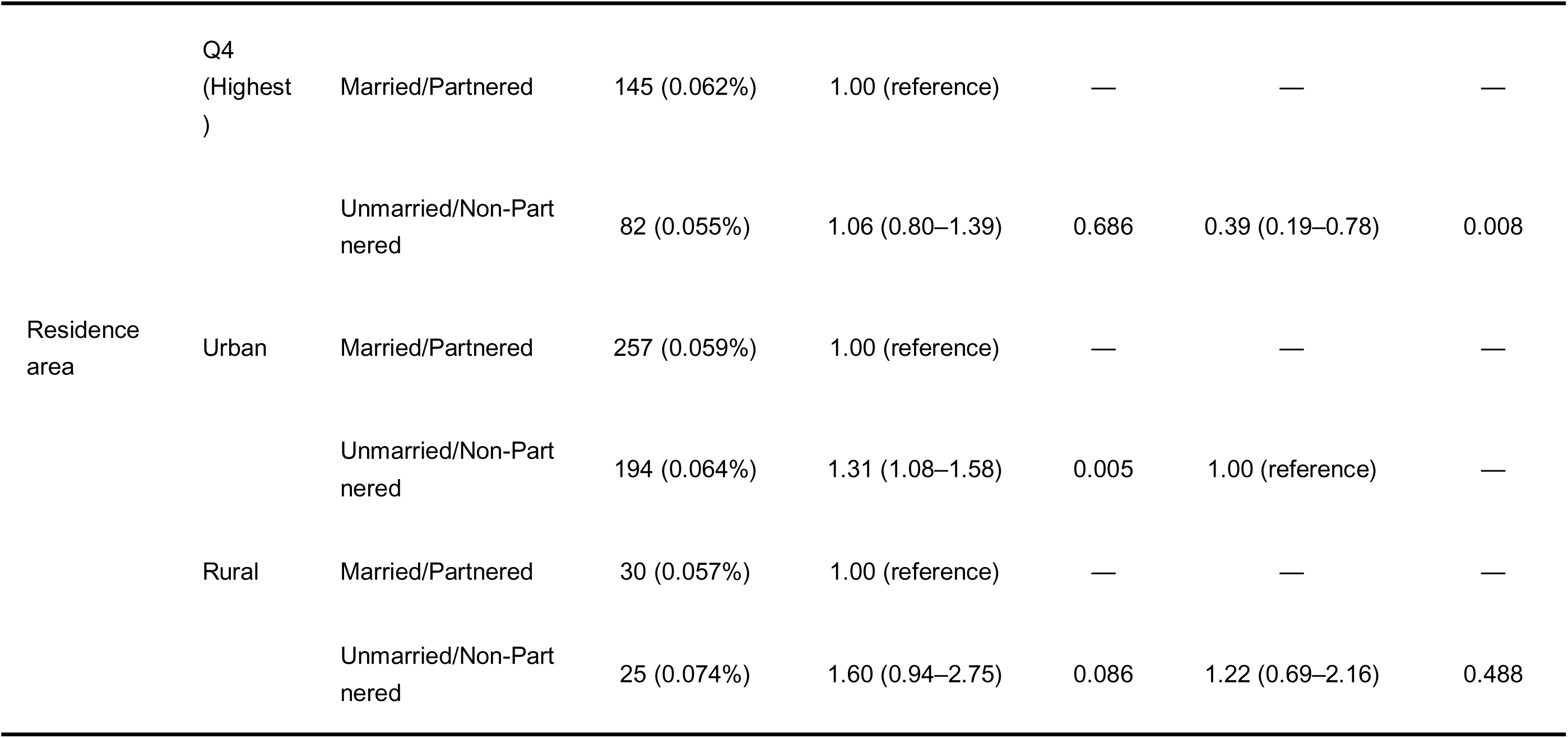
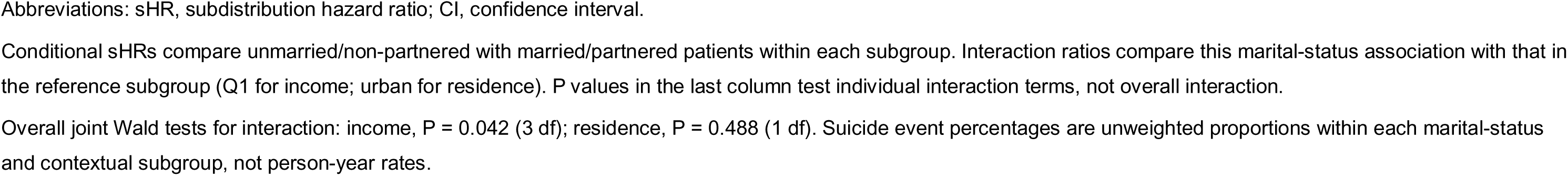
Stratum—Specific and Multiplicative Interaction Analysis of Marital Status on Suicide Mortality Moderated by Area—Level Median Household Income and. Rural-Urban Residence Area. Abbreviations: CI, confidence interval; sHR, subdistribution hazard ratio; sIPTW, stabilized inverse probability of treatment weighting; Q1-Q4, income categories 1 to 4. Stratum-specific conditional sHRs are relative subdistribution hazard ratios for unmarried/non-partnered versus married/partnered status within each income or residence category. They are obtained by adding the marital main-effect coefficient and applicable interaction coefficient on the log-sHR scale and exponentiating. Confidence intervals use the full robust covariance matrix, including coefficient covariance. Multiplicative interaction ratios compare that marital-status association with the Q1 or urban reference association. Overall interaction P values come from joint Wald tests (income: 3 df, P = 0.0424; residence: 1 df, P = 0.4877). Conditional effects are not absolute risk differences. Q1-Q4 denote fixed income categories.

### Subgroup analyses

Twelve of 22 subgroup levels yielded reportable estimates. Among patients aged <65 years, the marital-status sHR was 1.49 (95% CI 1.22-1.82), compared with 0.91 (0.62-1.34) among those aged _≥_65 years. Estimates were 1.46 (1.21-1.76) in White patients, 1.34 (1.06-1.69) for diagnosis in 2000-2010, 1.33 (1.01-1.77) for 2011-2022, 1.45 (1.15-1.82) for localized disease, and 1.29 (0.96-1.73) for regional disease. The Grade II estimate was 1.31 (95% CI 1.0005-1.7065; P = 0.0496). Chemotherapy-Yes and radiation-No/Unknown strata had sHRs of 1.43 (1.11-1.85) and 1.41 (1.10-1.80), respectively. Eight strata generated convergence warnings: Black race, Asian or Pacific Islander race, distant stage, Grades I and III, and all three surgery categories. American Indian/Alaska Native race and Grade IV had too few events for estimation. These unavailable estimates are not evidence of no association. Differences in subgroup-specific statistical significance do not establish effect modification (Figure 5).

**Figure 5:**
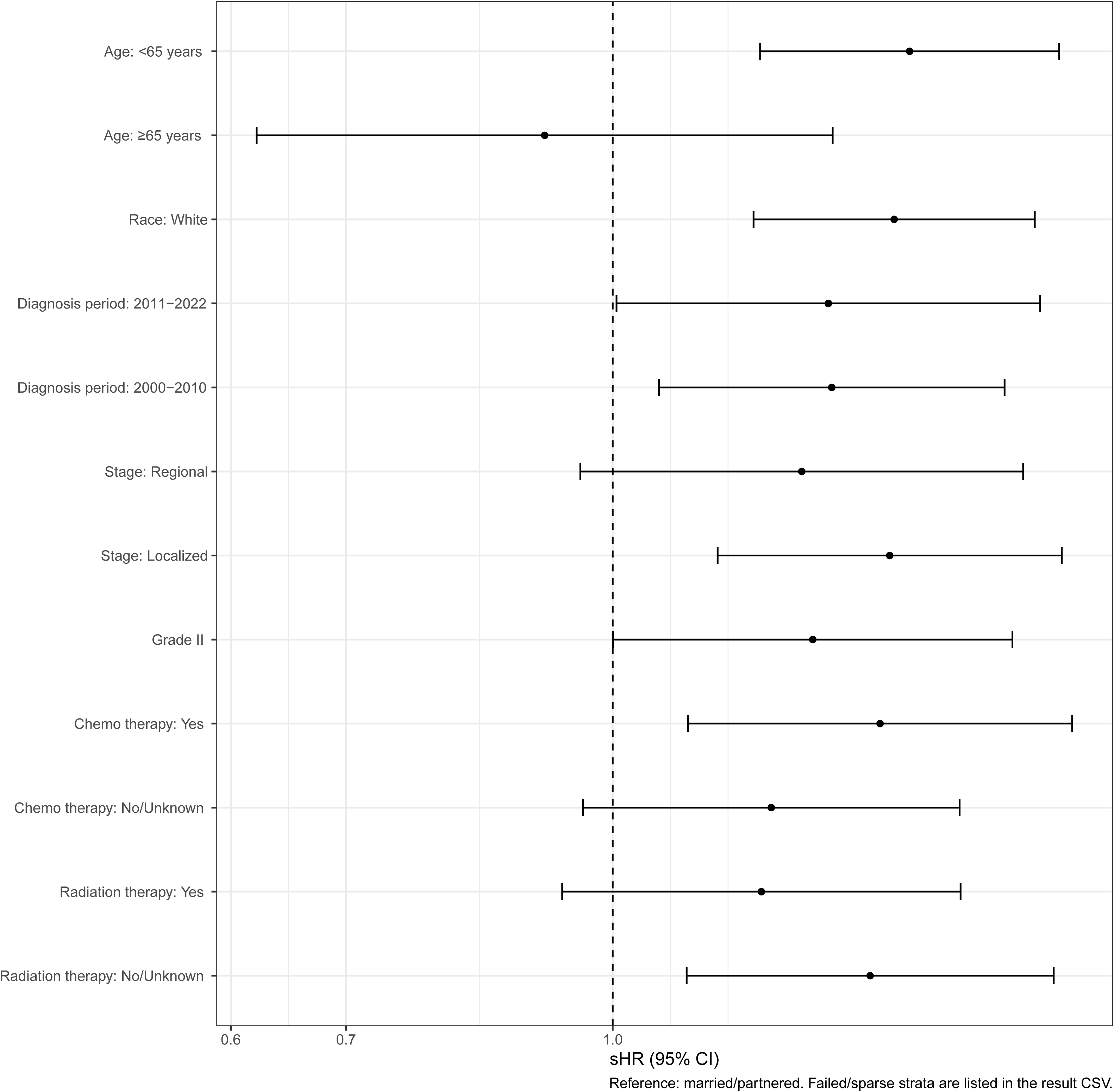
Subgroup analysis of marital status and suicide risk. Exploratory subgroup estimates use the baseline complete-case cohort, within-stratum Fine-Gray expansion, baseline sIPTW, remaining baseline covariates, and patient-clustered robust variance. Married/partnered is the reference. Twelve strata yielded estimates; eight had convergence warnings and two had insufficient events. Unavailable estimates are not statistically nonsignificant results. No formal tests comparing these subgroup effects were performed; unavailable strata are described in the Results.

### Sensitivity analyses

The unweighted treatment-adjusted analysis included 542,229 patients and 295 suicide deaths after excluding 282,818 patients (34.3% of the baseline complete-case cohort) whose surgery was classified as unknown. The marital-status estimate was attenuated and less precise than in the primary analysis (sHR 1.26, 95% CI 0.99-1.60; P = 0.0636; Figure 6). The confidence interval included 1. This analysis changed the sample, weighting, and covariate adjustment simultaneously and therefore does not isolate a treatment-adjustment effect.

**Figure 6:**
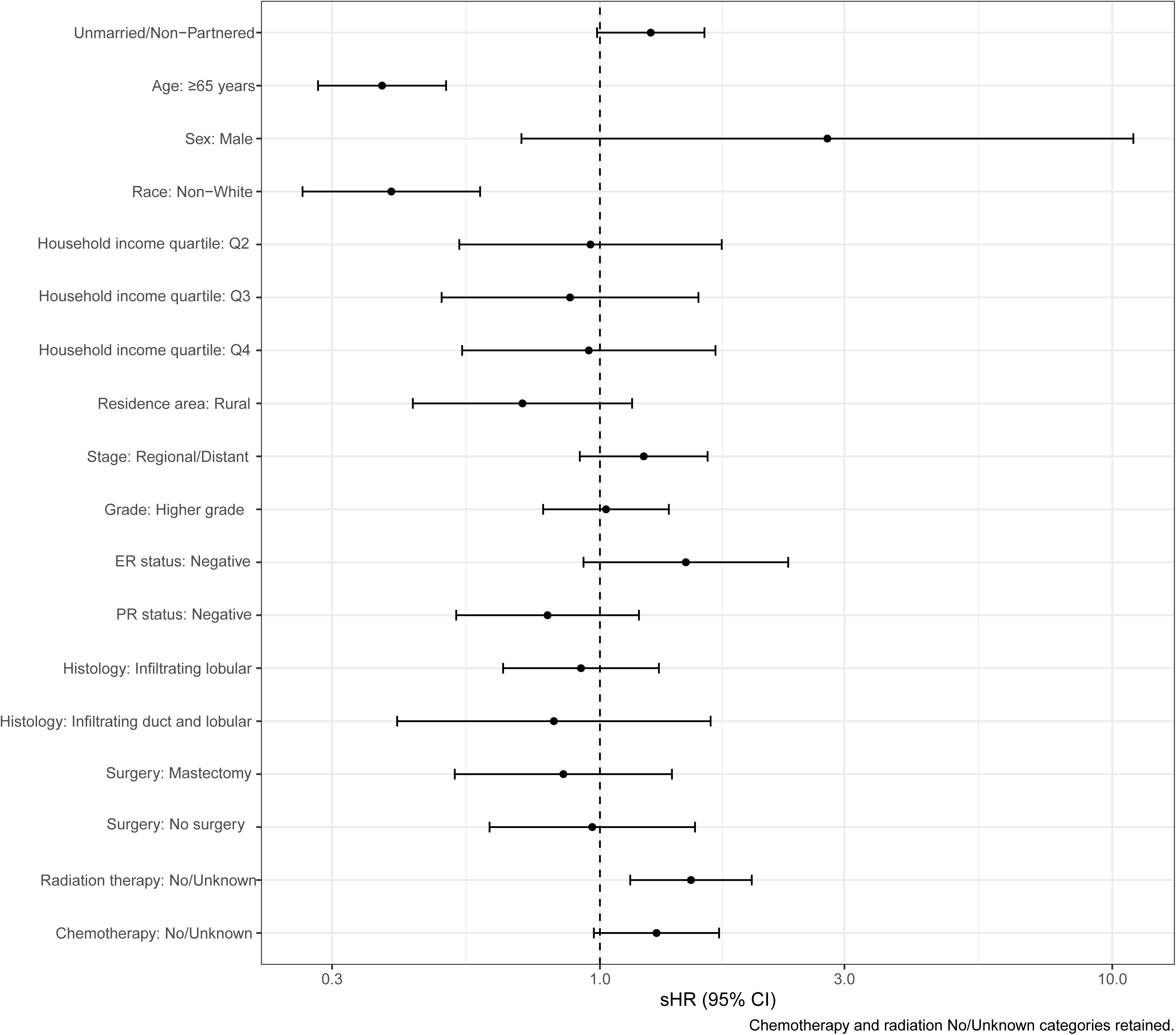
Sensitivity analysis. The unweighted treatment-adjusted Fine-Gray model included 542,229 patients and 295 suicide deaths, after excluding 282,818 records with surgery classified as unknown. The model adjusted for all primary baseline covariates, including sex, and for surgery, chemotherapy, and radiation. The combined chemotherapy and radiation No/Unknown categories were retained. sHRs and 95% CIs use patient-clustered robust variance. The marital-status sHR was 1.26 (95% CI 0.99-1.60; P = 0.0636). Treatment classification and selection limit interpretation.

## Discussion

In this population-based cohort, unmarried/non-partnered status was associated with higher suicide mortality in the primary Fine-Gray model, while the absolute rate was 7.42 per 100,000 person-years. Cox and landmark estimates had the same direction, but the treatment-adjusted sensitivity interval included the null. The exploratory income interaction suggested possible attenuation in higher-income counties; the Q4 conditional estimate was imprecise and not statistically significant. These findings concern associations with marital status at diagnosis and do not demonstrate that marriage itself protects against suicide.

The primary finding is consistent with earlier breast cancer research. Shi et al. reported higher suicide hazards for single and separated women than for married women in a competing-risk analysis (8). This is compatible with the Social Buffering Framework, but does not test its mechanisms (9). Marriage may correlate with emotional or practical support, while SEER does not measure relationship quality, caregiving, financial support, or help-seeking behavior. The association could also reflect selection and unmeasured differences in health or social circumstances.

The income interaction was exploratory. Although the joint test suggested heterogeneity, the Q4 conditional estimate did not exclude either a modest decrease or an increase in the relative subdistribution hazard. Suk et al. examined suicide after cancer diagnosis in relation to county-level income and rural-urban designation, providing a broader context for studying area-level conditions (28). Ayub et al. described rural-urban and income differences in breast cancer presentation and treatment rather than suicide outcomes (14). County income is an ecological measure and cannot be equated with an individual’s wealth, social support, or access to psycho-oncology care. Explanations involving substitution of community resources for spousal support remain hypotheses, and ecological fallacy is a concern.

The ER-negative association should also be interpreted cautiously. Shi et al. reported a higher suicide hazard for triple-negative than luminal B tumors, but receptor subtype contrasts are not equivalent to the ER-only comparison used here (8). That study observed 414 suicide deaths among 638,547 women; the present cohort had 506 events among 825,047 patients of both sexes. Differences in eligibility and observation periods preclude a direct comparison of risk. The small number of events relative to the number of model parameters limits precision, particularly in detailed tumor and treatment strata.

The current subgroup analysis did not produce a stable distant-stage estimate, so it cannot support the earlier claim of a lower marital-status sHR in distant disease. Competing non-suicide mortality may influence observed suicide incidence, but this mechanism was not demonstrated by the subgroup results. Sparse events, selection into survivor or treatment-defined strata, and model instability provide alternative explanations. Landmark analyses describe associations conditional on remaining alive and under observation; they do not eliminate survivor selection.

Marital status should not be used as a stand-alone suicide-screening criterion. Any clinical assessment should consider psychiatric history, current distress, social isolation, financial hardship, symptom burden, and access to care using established assessment approaches. Prior qualitative research has described support-related challenges among men with breast cancer (29), but the male main effect in this study does not show that the marital-status association is stronger in men: a marital-status-by-sex interaction was not tested. The present study did not evaluate a screening strategy or intervention.

Several limitations warrant consideration. Marital status was measured only at diagnosis and does not capture subsequent changes, relationship quality, or support networks; the unmarried group also combines heterogeneous circumstances. Psychiatric history and direct measures of social support were unavailable, leaving residual and unmeasured confounding. Exposure and cause-of-death misclassification are possible. Complete-case restriction, exclusion of the first month of observed survival, and treatment-related exclusions may introduce selection bias. County income is an ecological measure and cannot represent individual socioeconomic circumstances. Stage and treatment may partly reflect pathways linking social circumstances to outcomes, so the models do not estimate a total causal effect of marital status. Surgery classification and the large associated exclusions limit the treatment sensitivity analysis. Sparse events caused several subgroup fits to be non-estimable, and exploratory tests were not adjusted for multiplicity. The Fine-Gray proportional subdistribution hazards assumption was not formally assessed; stage and grade showed non-proportional hazards in the Cox model. The treatment-adjusted confidence interval included the null. These limitations constrain both causal and subgroup interpretations.

## Conclusion

Unmarried/non-partnered status at diagnosis was associated with a higher subdistribution hazard of suicide among patients with breast cancer, while the absolute suicide mortality rate remained low. Exploratory income interaction findings and the uncertain treatment-adjusted estimate require independent validation. The results support further investigation of social circumstances in survivorship without establishing a causal effect of marriage or a stand-alone screening rule.

## Data Availability

All data produced in the present study are available upon reasonable request to the authors.

## Abbreviations

sHR: Subdistribution Hazard Ratio;
CI: Confidence Interval;
SEER: Surveillance, Epidemiology, and End Results;
sIPTW: Stabilized Inverse Probability of Treatment Weighting;
SMD: Standardized Mean Difference;
Cox: Cox Proportional Hazards;
ER: Estrogen Receptor;
PR: Progesterone Receptor;
HR: Hazard Ratio;
aHR: Adjusted Hazard Ratio;
Q1-Q4: Income categories 1 to 4;
CIF: Cumulative Incidence Function;
P: P-value;
SMR: Standardized Mortality Ratio.

## Author Contributions

Xuehai Zou had full access to all of the data in the study and takes responsibility for the integrity of the data and the accuracy of the data analysis.

*Concept and design:* Jingyi Shi, Xuehai Zou.

*Acquisition, analysis, or interpretation of data:* Xuehai Zou.

*Drafting of the manuscript:* Jingyi Shi, Xuehai Zou.

*Critical review of the manuscript for important intellectual content:* Jingyi Shi, Xuehai Zou.

*Statistical analysis:* Xuehai Zou.

*Supervision:* Jingyi Shi.

## Acknowledgements

Not applicable.

## Ethics Statement

This study analyzed de-identified SEER registry data under the applicable data-use agreement. It involved no direct participant contact or collection of identifiable information.

## Data Availability

SEER data can be requested from the US National Cancer Institute through the applicable registration and data-use agreement process. Patient-level records cannot be redistributed by the authors. Analysis code, variable definitions, and permitted aggregate outputs may be made available in accordance with the SEER agreement and journal policy.

---

**Supplementary Figure 1:**
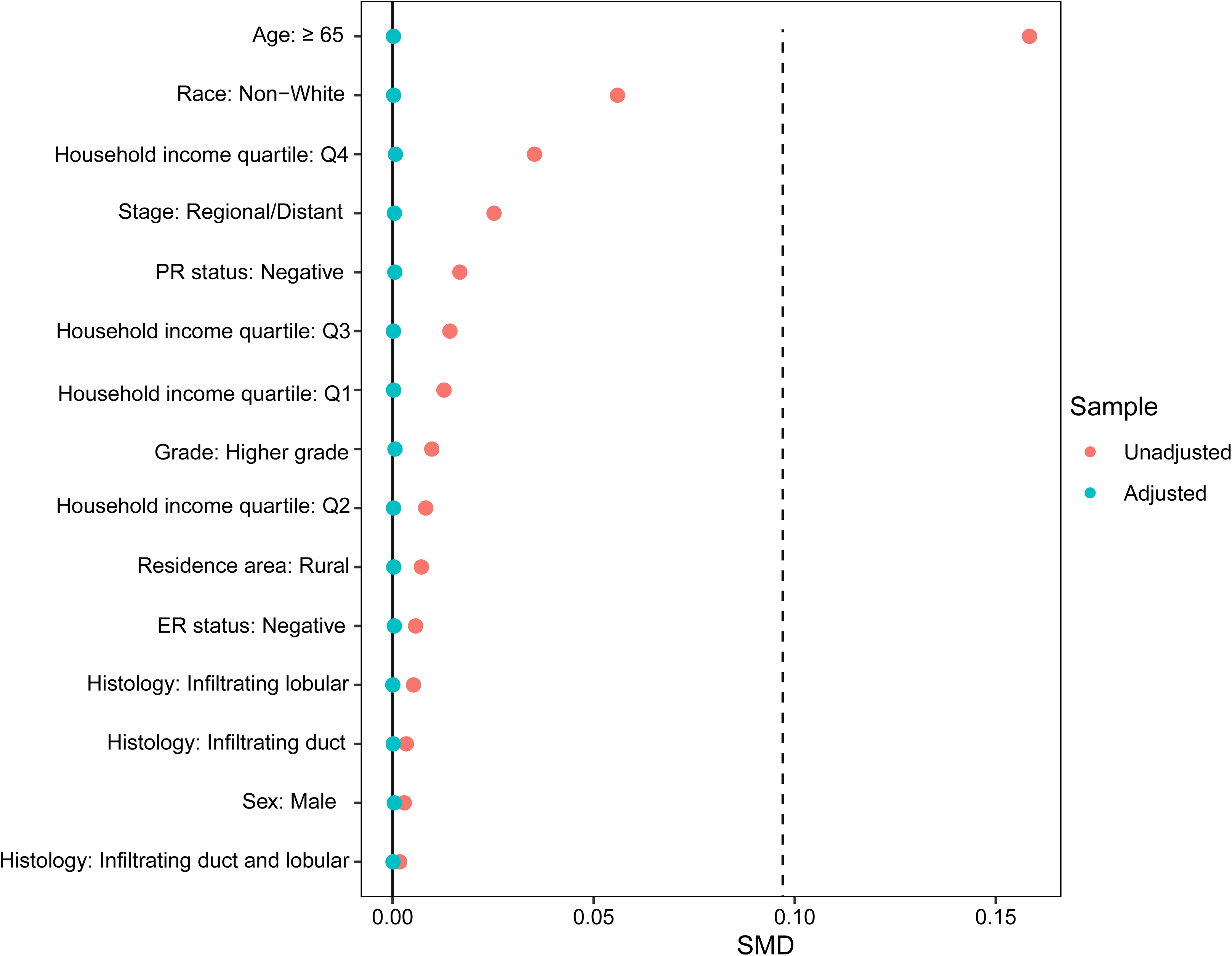
Covariate balance before and after sIPTW. Covariate balance before and after sIPTW is displayed for the baseline variables. Standardized mean differences are reported in Table 1, where all post-weighting SMDs were below 0.01 (maximum 0.00536). The separately submitted plot and the tabulated SMDs are complementary balance summaries.

**Supplementary Figure 2:**
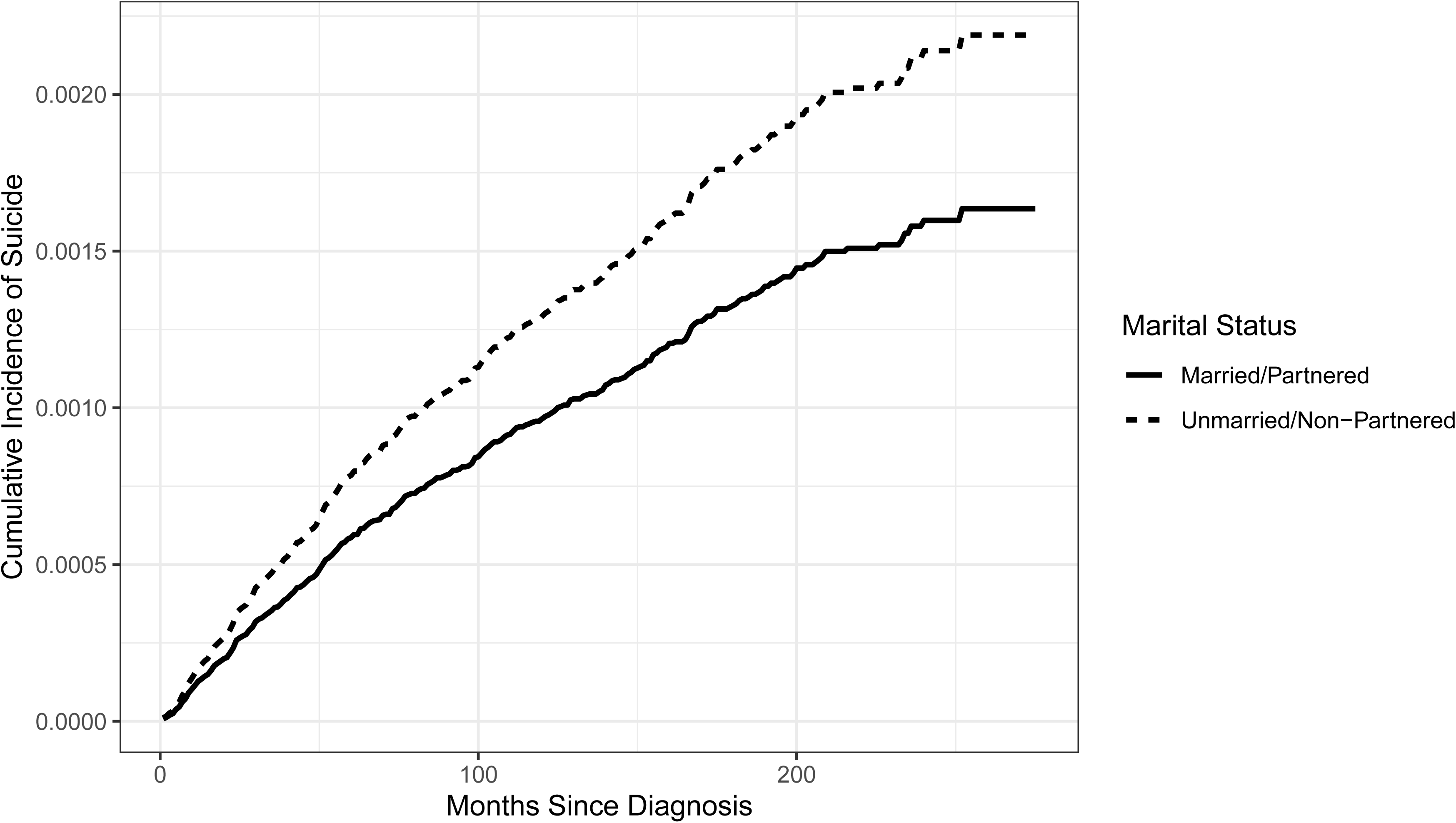
Fine-Gray cumulative incidence of suicide by marital status. Cumulative incidence curves were predicted from the sIPTW-weighted multivariable Fine-Gray model for two otherwise identical reference profiles: age <65 years, female, White, Q1 income, urban residence, localized stage, lower grade, ER positive, PR positive, and infiltrating ductal histology. Only marital status differs. These are reference-profile predictions, not population-standardized CIFs or crude observed mortality rates. Time is measured in months since diagnosis.

**Supplementary Table 1:** Patient Characteristics Before Weighting by Marital Status. Abbreviations: SD, standard deviation; ER, estrogen receptor; PR, progesterone receptor. This table describes the same 825,047-patient baseline complete-case cohort as Table 1 before weighting. Continuous age is shown as mean (SD), and categorical characteristics as n (%). Treatment variables are descriptive and were not required for the primary complete-case cohort; standalone unknown surgery and combined chemotherapy/radiation No/Unknown categories are retained. Descriptive P values are from the supplied table output and do not establish clinical importance. Exposure, outcome, baseline coding, and income cutoffs are defined in the Methods.

| Characteristic | Overall N =<br>825,047<sup>1</sup> | Married/Partnered N =<br>489,169<sup>1</sup> | Unmarried/Non-Partnered N = 335,878<sup>1</sup> | p-value<sup>2</sup> |
| --- | --- | --- | --- | --- |
| **Age at Diagnosis** | 60.2 (13.5) | 58.2 (12.3) | 63.1 (14.5) | <0.001 |
| **Age** |  |  |  | <0.001 |
| <65 | 508,122<br>(62%) | 332,807 (68%) | 175,315 (52%) |  |
| ≥65 | 316,925<br>(38%) | 156,362 (32%) | 160,563 (48%) |  |
| **Sex** |  |  |  | <0.001 |
| Female | 819,607<br>(99%) | 485,362 (99%) | 334,245 (100%) |  |
| Male | 5,440 (0.7%) | 3,807 (0.8%) | 1,633 (0.5%) |  |
| **Race** |  |  |  | <0.001 |
| American<br>Indian/Alaska Native | 4,364 (0.5%) | 2,267 (0.5%) | 2,097 (0.6%) |  |
| Asian or Pacific<br>Islander | 76,520<br>(9.3%) | 52,564 (11%) | 23,956 (7.1%) |  |
| Black | 83,220<br>(10%) | 31,328 (6.4%) | 51,892 (15%) |  |
| White | 660,943<br>(80%) | 403,010 (82%) | 257,933 (77%) |  |
| **Household Income**<br>**Quartile** |  |  |  | <0.001 |
| Q1 | 59,719<br>(7.2%) | 32,862 (6.7%) | 26,857 (8.0%) |  |
| Q2 | 130,655<br>(16%) | 75,824 (16%) | 54,831 (16%) |  |
| Q3 | 254,131<br>(31%) | 147,832 (30%) | 106,299 (32%) |  |
| Q4 | 380,542<br>(46%) | 232,651 (48%) | 147,891 (44%) |  |
| **Residence Area** |  |  |  | <0.001 |
| Urban | 738,862<br>(90%) | 436,645 (89%) | 302,217 (90%) |  |
| Rural | 86,185<br>(10%) | 52,524 (11%) | 33,661 (10%) |  |
| **Diagnosis Period** |  |  |  | <0.001 |
| 2000-2004 | 141,789<br>(17%) | 84,699 (17%) | 57,090 (17%) |  |
| 2005-2009 | 171,690<br>(21%) | 101,287 (21%) | 70,403 (21%) |  |
| 2010-2014 | 183,985<br>(22%) | 108,487 (22%) | 75,498 (22%) |  |
| 2015-2019 | 205,434<br>(25%) | 121,889 (25%) | 83,545 (25%) |  |
| 2020-2022 | 122,149<br>(15%) | 72,807 (15%) | 49,342 (15%) |  |
| **Stage** |  |  |  | <0.001 |
| Localized | 524,213<br>(64%) | 315,829 (65%) | 208,384 (62%) |  |
| Regional | 266,208<br>(32%) | 156,761 (32%) | 109,447 (33%) |  |
| Distant | 34,626<br>(4.2%) | 16,579 (3.4%) | 18,047 (5.4%) |  |
| **Grade** |  |  |  | <0.001 |
| Grade I | 183,689<br>(22%) | 111,184 (23%) | 72,505 (22%) |  |
| Grade II | 371,613<br>(45%) | 219,994 (45%) | 151,619 (45%) |  |
| Grade III | 265,291<br>(32%) | 155,291 (32%) | 110,000 (33%) |  |
| Grade IV | 4,454 (0.5%) | 2,700 (0.6%) | 1,754 (0.5%) |  |
| **ER Status** |  |  |  | <0.001 |
| Positive | 678,126<br>(82%) | 403,200 (82%) | 274,926 (82%) |  |
| Negative | 146,921<br>(18%) | 85,969 (18%) | 60,952 (18%) |  |
| **PR Status** |  |  |  | <0.001 |
| Positive | 590,809<br>(72%) | 353,620 (72%) | 237,189 (71%) |  |
| Negative | 234,238<br>(28%) | 135,549 (28%) | 98,689 (29%) |  |
| **Histology** |  |  |  | <0.001 |
| Infiltrating duct | 675,010<br>(82%) | 399,531 (82%) | 275,479 (82%) |  |
| Infiltrating lobular | 126,358<br>(15%) | 75,954 (16%) | 50,404 (15%) |  |
| Infiltrating duct and<br>lobular | 23,679<br>(2.9%) | 13,684 (2.8%) | 9,995 (3.0%) |  |
| **Surgery** |  |  |  | <0.001 |
| Breast-conserving<br>surgery | 461,643<br>(56%) | 278,743 (57%) | 182,900 (54%) |  |
| Mastectomy | 35,110<br>(4.3%) | 21,125 (4.3%) | 13,985 (4.2%) |  |
| No surgery | 45,476<br>(5.5%) | 19,471 (4.0%) | 26,005 (7.7%) |  |
| Unknown | 282,818<br>(34%) | 169,830 (35%) | 112,988 (34%) |  |
| **Chemotherapy** |  |  |  | <0.001 |
| Yes | 359,300<br>(44%) | 226,407 (46%) | 132,893 (40%) |  |
| No/Unknown | 465,747<br>(56%) | 262,762 (54%) | 202,985 (60%) |  |
| **Radiation Therapy** |  |  |  | <0.001 |
| Yes | 456,105<br>(55%) | 284,468 (58%) | 171,637 (51%) |  |
| No/Unknown | 368,942<br>(45%) | 204,701 (42%) | 164,241 (49%) |  |

<sup>1</sup>Mean (SD); n (%)
<sup>2</sup>Wilcoxon rank sum test; Pearson's Chi-squared test

